# Characteristics of patients with positional OSA according to ethnicity and the identification of a novel phenotype – Lateral Positional Patients (Lateral PP): A MESA study

**DOI:** 10.1101/2022.05.03.22274534

**Authors:** Yuval Ben Sason, Arie Oksenberg, Jonathan A. Sobel, Joachim A. Behar

## Abstract

**Study Objectives:** To investigate the characteristics of Obstructive Sleep Apnea (OSA) positional patients’ (PP) phenotypes among different ethnic groups in the Multi-Ethnic Study of Atherosclerosis (MESA) dataset. Moreover, we hypothesized the existence of a new OSA PP phenotype we coined “Lateral PP”, for whom the lateral apneas hypopnea index (AHI) is at least double the supine AHI.

**Methods:** From 2,273 adults with sleep information, we analyzed data of 1,323 subjects that slept more than 4 hours and had at least 30 minutes of sleep in both the supine and the non-supine positions. Demographics and clinical information were compared for the different PP, and ethnic groups.

**Results:** 861 (65.1%) patients had OSA and 35 (4.1%) were Lateral PP. Lateral PP patients were mainly females (62.9%), obese (31.4 median body mass index), had mild to moderate OSA (94.3%), and mostly were non-Chinese American (97.1%). Among all OSA patients, 550 (63.9%) were Supine PP, and 17.7% were supine-isolated OSA (siOSA). Supine PP and Lateral PP were present in 73.1% and 1.0% of Chinese Americans, 61.0% and 3.4% of Hispanics, 68.3% and 4.7% of White-Caucasian, and 56.2% and 5.2% of Black-African American OSA patients.

**Conclusion:** Chinese-American have the highest prevalence of Supine PP, whereas Black-African American patients lean towards less Supine PP and higher Lateral PP. Lateral PP appears as a novel OSA phenotype. However, it was found for a small group of OSA patients and thus its presence should be further validated.

**Brief Summary:** We studied prevalence of obstructive sleep apnea (OSA) positional phenotypes among Black African-American, Caucasian, Hispanic, and Chinese American patients and described their demographic and polysomnographic characteristics. Despite similar levels of OSA severity in the four ethnic groups, Chinese American had a higher prevalence of Supine PP, whereas Black-African American patients were significantly less Supine PP, as compared to other ethnic groups.

In addition, we identify a novel OSA PP phenotype we named “Lateral positional patients (Lateral-PP)”. These OSA patients had apnea and hypopnea events mainly in the lateral position. This was a small group of OSA patients (4.1%) that were mainly obese, female, with mild-moderate OSA, and more prevalent in Black-African American.

## Introduction

Obstructive sleep apnea (OSA) is characterized by periods of breathing cessation (apnea) and of reduced breathing effort (hypopnea) during sleep due to the complete or partial collapse of the upper airway. OSA is estimated to affect 9-38% of the adult population, and even more among elderly people^1^. Today, the medical definition of OSA is actively being challenged because of the different characteristics of OSA individuals. A good example of different phenotypes is OSA Positional Patients (PP) vs. Non-Positional Patients (NPP). PP have sleep-related breathing abnormalities mainly when they sleep in the supine posture. Those are patients with OSA that can benefit from Positional Therapy (PT), i.e., the avoidance of the supine posture during sleep. On the contrary, PT for NPP could only partially help this OSA group^2^. Thus, different phenotypes of OSA should be diagnosed and managed in different manners. It is thus critical to define these different phenotypes of the disease in order to personalize the diagnosis and treatment pathway for patient presenting OSA. OSA is divided into three severity levels according to the apnea-hypopnea index (AHI) which corresponds to the average number of apnea or hypopnea events per hour; mild OSA with an AHI greater than 5 and lower than 15, moderate OSA with an AHI in the range of 15 to 30, and severe OSA with an AHI above 30^3^.

Many researches have shown the presence of OSA phenotypes in which symptoms appearance and severity are highly related to sleep positions^4–9^. The pioneer study of Cartwright RD in 1984^9^ showed the worsening effect of the supine posture on the Apnea Index in 30 patients with OSA. Oksenberg et al., 1997^8^ described the demographic and polysomnographic characteristics of PP vs. NPP, and showed that PP are a dominant group of OSA patients representing 55.9% of all patients with OSA evaluated in a Sleep Unit. This study also showed that PP are more frequent in Mild-Moderate OSA (ranging from 65-69%), and that PP are typically thinner and younger, have less severe breathing abnormalities and suffer less from daytime sleepiness than NPP. Later, it was found that Asian patients with OSA have a much higher prevalence of PP reaching 75% of all OSA and up to 87% in mild OSA^10^. Thus PT could represent a simple, low cost and effective therapy to minimize the healthy burden of many patients with mild-moderate OSA^11^. It is of interest that it has not been investigated how PP varies in different ethnic OSA groups, therefore we analyzed the prevalence and characteristics of PP according to the ethnicity in a relatively large group of OSA patients. In addition, we hypothesized the existence of a group of OSA patients for whom the Lateral AHI can be at least double the Supine AHI and investigate the prevalence and characteristics of this novel PP phenotype that we coined “Lateral PP”.

## Methods

### Multi-ethnic study of Atherosclerosis

A polysomnography (PSG) database of 2,273 adults (age≥54), collected between November 2010 to 2012 by the National Heart, Lung and Blood Institute (NHLBI) as a part of the Multi-Ethnic Study of Atherosclerosis (MESA)^12^. The data was collected at-home and included a variety of ethnicities; Black or African-American, White or Caucasian, Hispanic, and Chinese American men and women. This dataset is available from the National Sleep Research Resources (NSRR) and denoted as MESA database^13^. A total of 2,056 had raw PSG data available, among them 1,323 recordings had minimum of 4 hours total sleep time (TST), and at least 30 min spent in both the supine and non-supine position, were included in the analysis. PSG recordings included brain waves, electrocardiogram, eye-movement, body position, blood oxygen level, breathing pattern, chest movement, abdominal movement and snoring during sleep. Permission to use retrospective medical databases was granted following internal institutional review board (IRB; #62-2019).

### Respiratory events’ definitions

To calculate the AHI, obstructive apnea, and recommended hypopnea events were used. An obstructive apnea event was defined by a drop of 90% or more in the amplitude of the airflow signal from a baseline period (i.e. a regular breathing period with stable oxygen levels breathing). This needed to hold for at least 90% of the event duration, and for a minimum event duration of 10 seconds. A recommended hypopnea event was defined as a reduction from a baseline of > 70% in breathing amplitude that lasted 10 seconds or more. Hypopneas events had to be associated with a 4% oxygen desaturation.

### PP and its derivatives’ definitions

In relation to positionality, we divided sleep MESA database into the following four groups (Figure 1) –

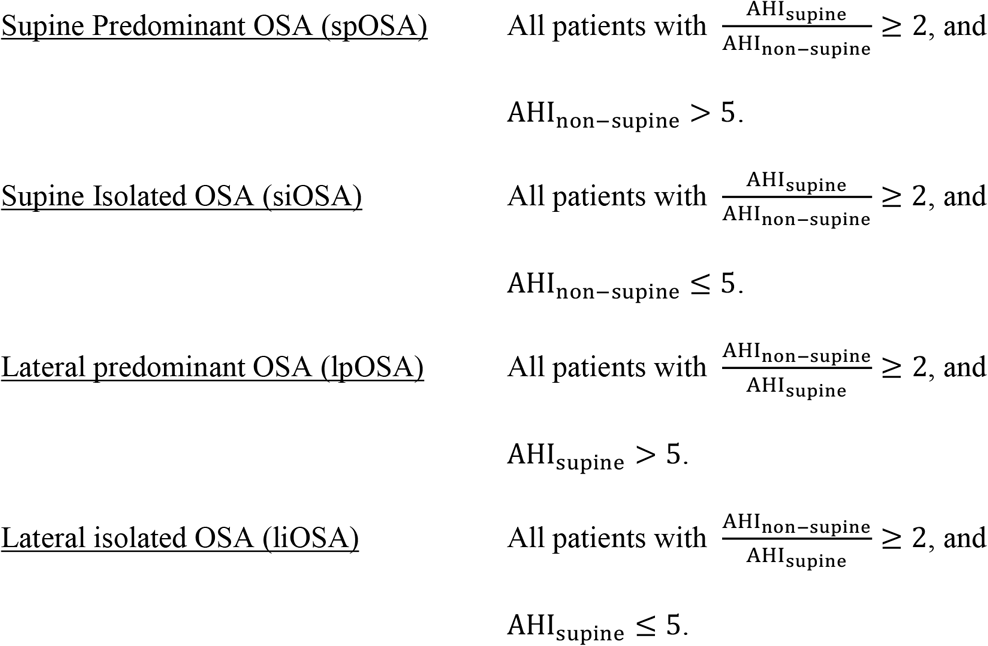

**Figure 1:**
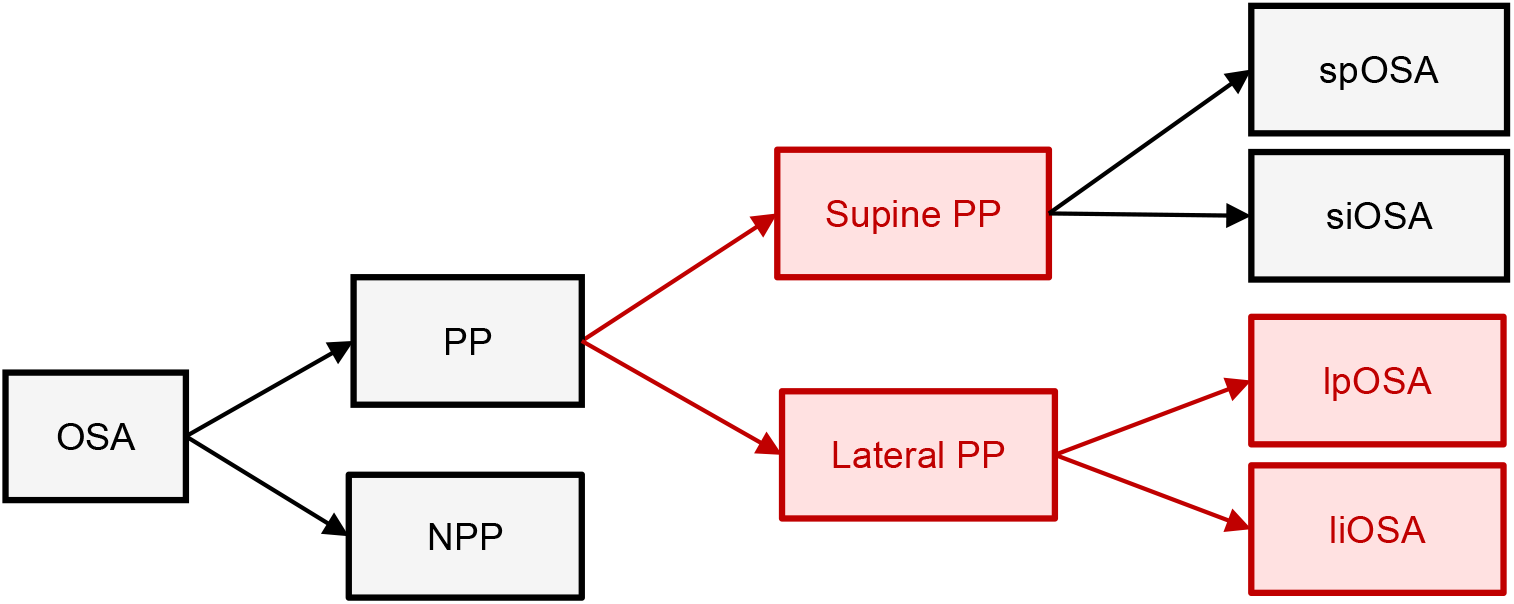
OSA phenotyping flowchart illustration. OSA and its derivatives’ diagram according to positional effect. In red is the new Positional Patients (PP) suggested phenotype and its sub-phenotypes. sp = Supine predominant, si = Supine isolated, lp = Lateral predominant, li = Lateral isolated. NPP = Non-positional patients.

As showed in Figure 1, Lateral PP include both lpOSA and liOSA, as well as Supine PP include both spOSA and siOSA.

### Statistical tests

We used Kolmogorov-Smirnov (KS) test to compare the distribution of TST, TST in supine position (TST_supine_), TST in non-supine positions (TST_non−supine_), and the ratio of TST_supine_ by the 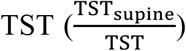, between the Lateral PP groups, and all the other patients. The Mann-Whitney nonparametric statistical test (U-test) was used to assess the assumption that AHI_non−supine_ of Lateral PP, is greater than the AHI_non−supine_ of the supine PP. For the multivariate test, to compare between the characteristics of the different OSA phenotypes, or different ethnic groups, we used Kruskal-Wallis (KW) H-test for independent samples. We also used this statistical test to examine the prevalence of different OSA phenotypes among patients with OSA from different ethnicities. Statistical tests were made in Python 3 using the statistical functions (‘stats’) of the SciPy 1.6.1 package. A multivariate logistic model was performed using the generalized linear model framework in R, to model PP versus NPP, and Lateral PP versus Supine PP, based on BMI, gender, age, race and co-morbidities. Statistical significance was considered for both-sided p-value ≤ 0.05 and odd ratio (OR) were reported. Summary statistics were reported as median and interquartile range (IQR).

## Results

### OSA severity groups and PP prevalence

861 of these patients were OSA patients with overall AHI ≥ 5/*h*. Their continuous and categorical demographical, clinical and polysomnographic data are summarized in Table 1. Figure 2 summarizes how the patients included in this study are divided. A total number of 178 patients slept less than 4 hours, and 541 did not sleep for a minimum of 30 minutes in supine, and/or in non-supine positions. An additional 14 patients had no documented AHI (i.e., AHI=None), and 462 patients had AHI ≤ 5/*h*, and were considered as non-OSA. Supine Positional Patients (Supine PP), following Cartwright definition^9^, was found in 550 (63.9%) out of 861 OSA patients. Supine PP was found in 306 of 443 patients (69.07%) with mild OSA, 158 of 254 patients (62.20%) with moderate OSA, and 86 of 164 patients (52.44%) with severe OSA. The total number of patients with siOSA were 232 with mild OSA (75.82% of the mild Supine PP, and 52.37% of the total number of mild OSA), 48 with moderate OSA (30.38% of the moderate Supine PP, and 18.90% of the total number of moderate OSA) and 13 with severe OSA (15.12% of the severe Supine PP, and 7.93% of the total number of severe OSA). Figure S2 visualize the fraction of Supine PP, siOSA, spOSA and NPP for each OSA severity group, and also the fraction of spOSA, and siOSA in each severity’s Supine PP group.

**Table 1:**
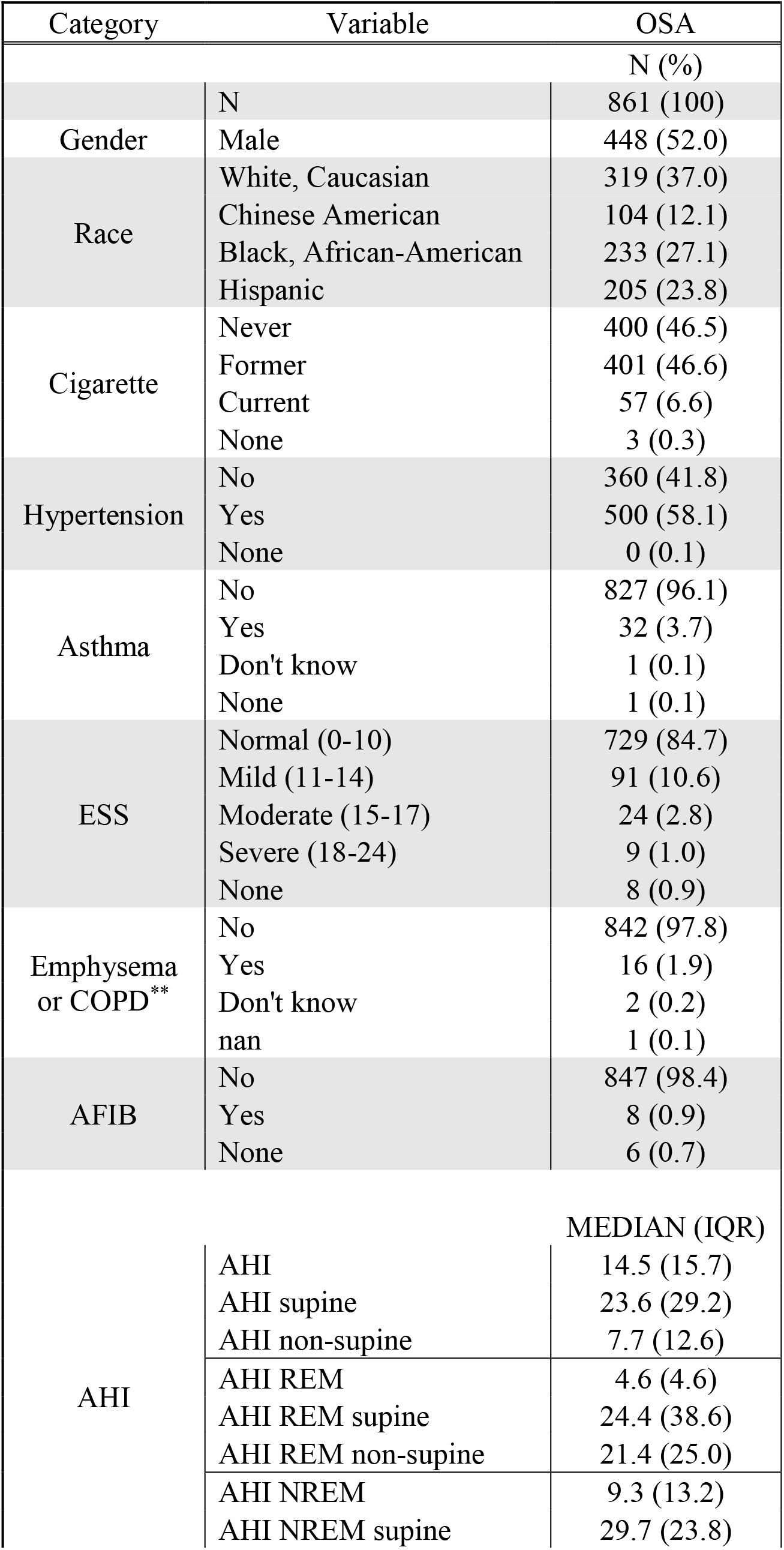
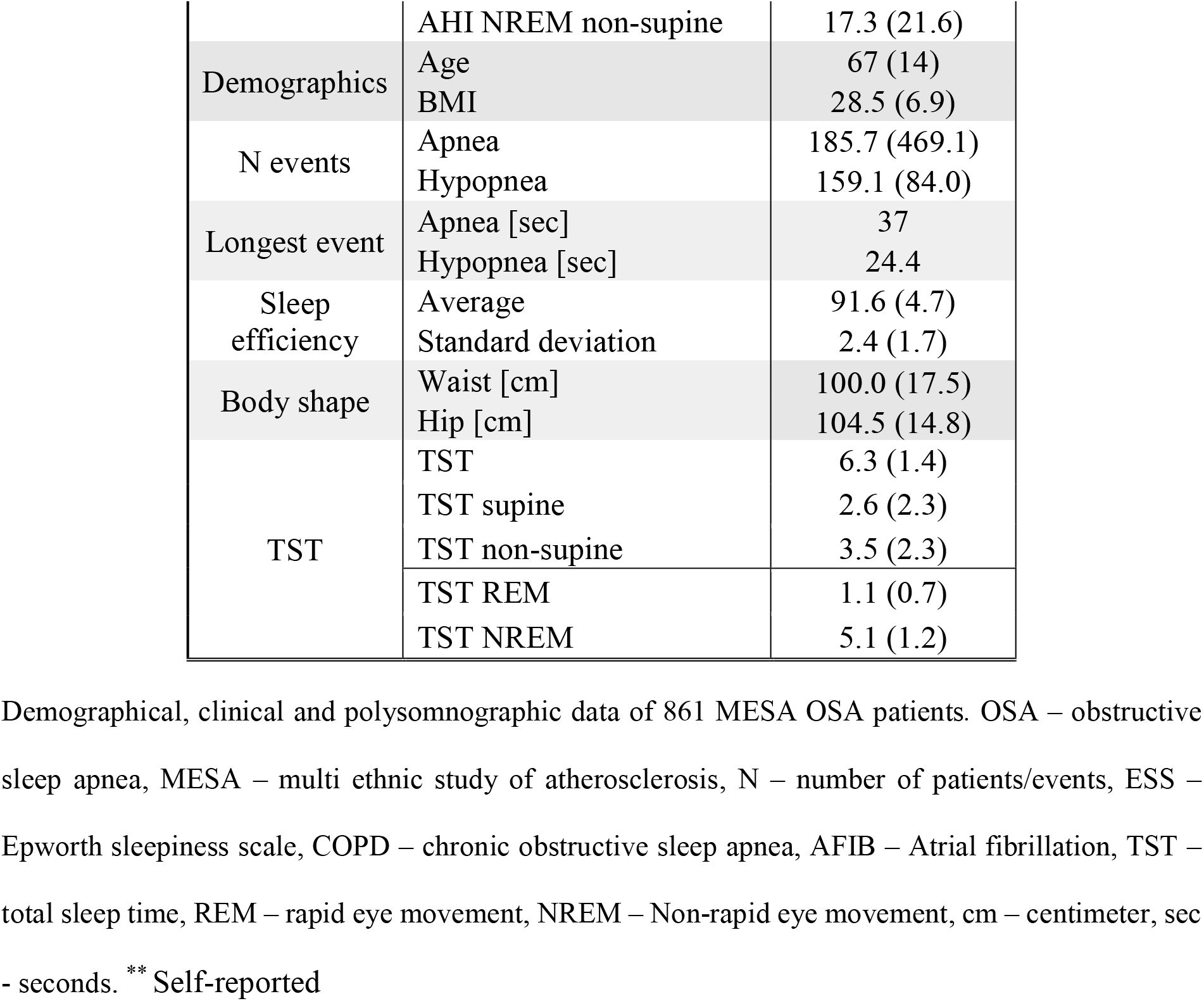
Demographical, clinical and polysomnographic data of 861 OSA patients.

**Figure 2:**
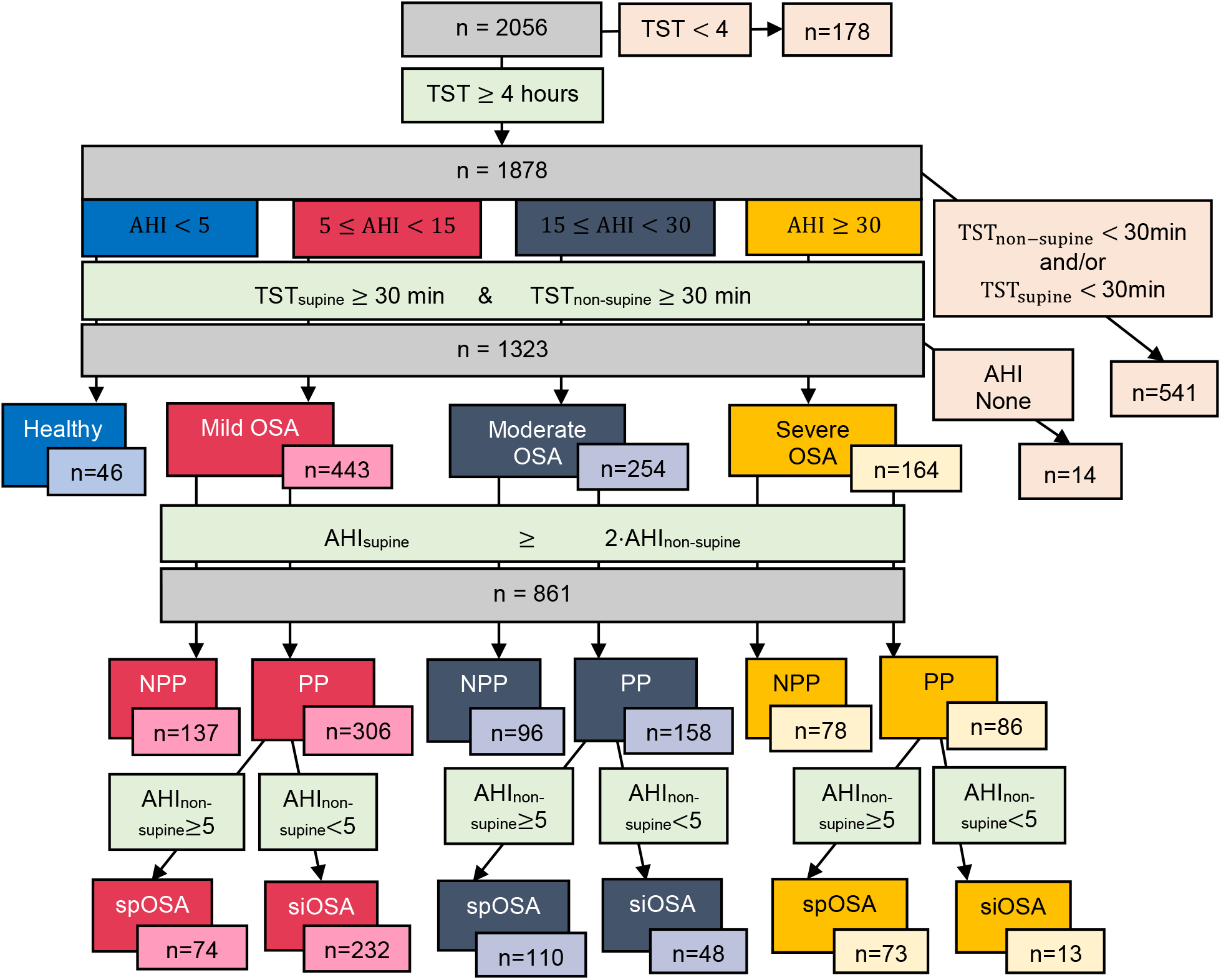
MESA patients division per condition and per exclusion criteria. Block diagram of MESA patients and exclusion criteria. AHI – Apnea Hypopnea Index, OSA – Obstructive Sleep Apnea, PP – Positional Patients, NPP – Non PP, spOSA – Supine Predominant OSA, siOSA – Supine Isolated OSA, n – Number of patients, min - minutes.

### Lateral Positional Patients (Lateral PP) analysis

Among the 861 OSA subjects, 35 patients (4.1%) were Lateral PP with 14 lpOSA and 21 liOSA. From these 35 patients, 12 patients slept less than 1 hour in either supine or non-supine position (10 in supine, 2 in non-supine), as seen in Table. According two sample KS statistical test, the distributions of the TST_supine_, TST_non−supine_, and TST_supine_/TST, per Lateral and Supine PP groups, were significantly different (p-values < 0.05).

shows that most of the TST of the Lateral PP group was in the lateral position with only 4 out of 35 patients having TST_prone_ greater than 30 minutes and still with a meaningful amount of TST_lateral_ (between 1.48-2.85 hours). Figure S3 and Figure S4 show the distributions of the sleep times for the different sleeping position and for the Lateral PP group, and Supine PP group. The non-supine AHI was significantly higher (p-value < 0.001) for Lateral PP (13.7 (13.3)) than for Supine PP (4.5 (7.6)).

Referring to Table 2, the Lateral PP were mainly obese (31.4 (4.9) BMI), with significantly higher (p-value < 0.005) BMI than patients with spOSA (29.0 (5.9)) and patients with siOSA (26.9 (5.2)). They were mainly female and had a significantly higher proportion than spOSA female prevalence (62.9% versus 39.3%, p-value= 0.008). They were also patients with mild-moderate OSA (94.3%), and were mostly non-Chinese American (97.1%), as can be seen in Table 3. In addition, most of them were hypertensive (65.7%), former smoker (51.4%), and did not suffer from excessive daytime sleepiness according to the Epworth Sleepiness Scale (ESS) (88.4%). Though, for these three last parameters, the prevalence was not significantly different from the other OSA phenotypes. Twenty-one patients out of the 35 (60.0%) were liOSA but nine of them (42.9%) slept less than 1 hour in the supine posture and the 15 remaining slept less than 2.05 hours in this posture. Out of the 35 Lateral PP, 10 slept less than 1-hour in supine position and 2 slept less than 1 hour in the lateral position. Still, it is important to notice that 25 out of the 35 Lateral PP slept more than 1 hour in the supine posture. The multivariate logistic model between PP and NPP showed that age and BMI were significant predictors of NPP with ORs of 1.02 (p-value = 0.004) and 1.1 (p-value = 0.017) respectively. This suggests that NPP are older and over-weighted. In addition, the multivariate logistic model between Supine PP and Lateral PP revealed that BMI was a significant predictor of Lateral PP, with an OR of 1.25 (p-value = 0.019), suggesting that Lateral PP are more obese patients.

**Table 2:**
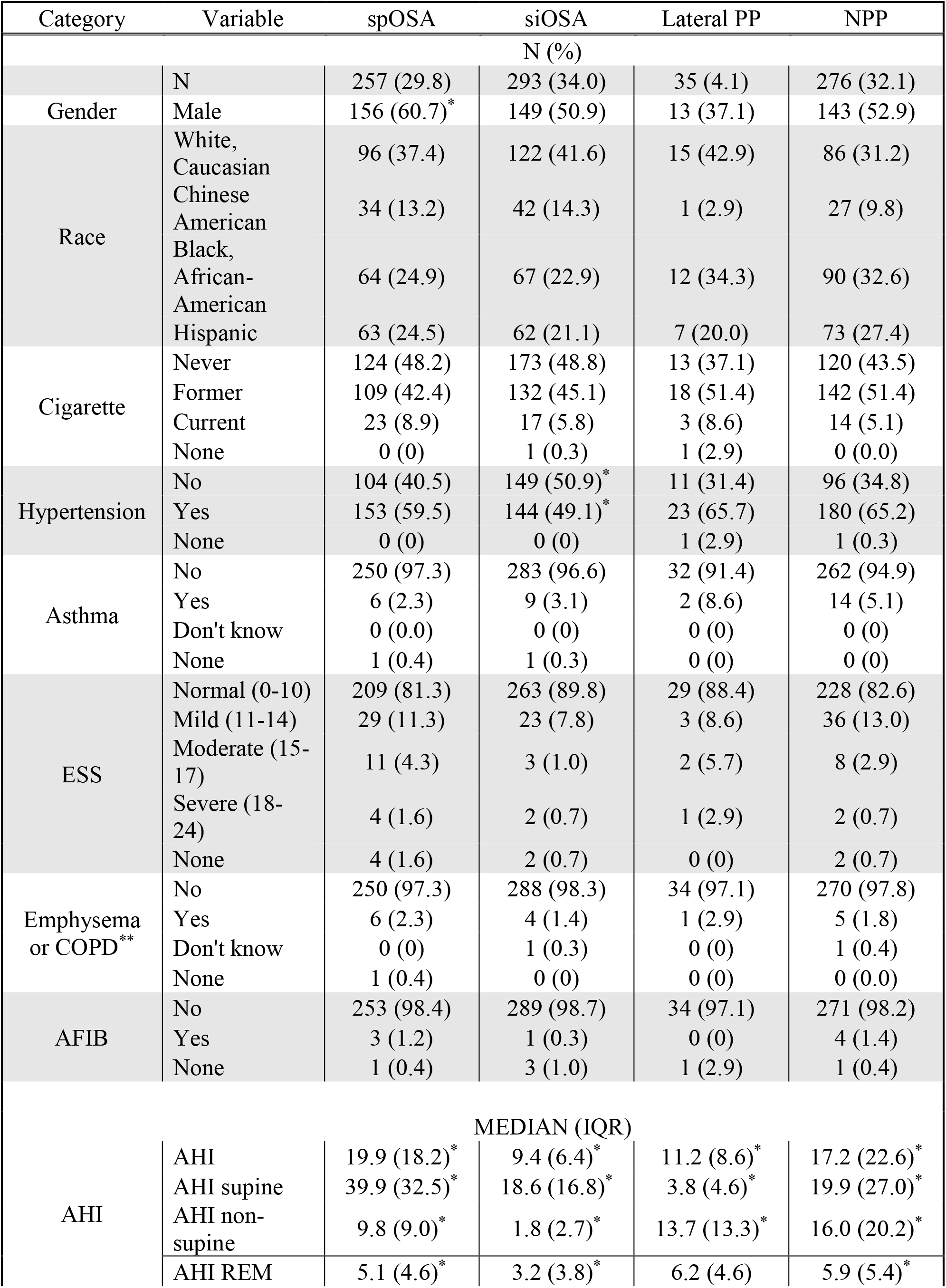
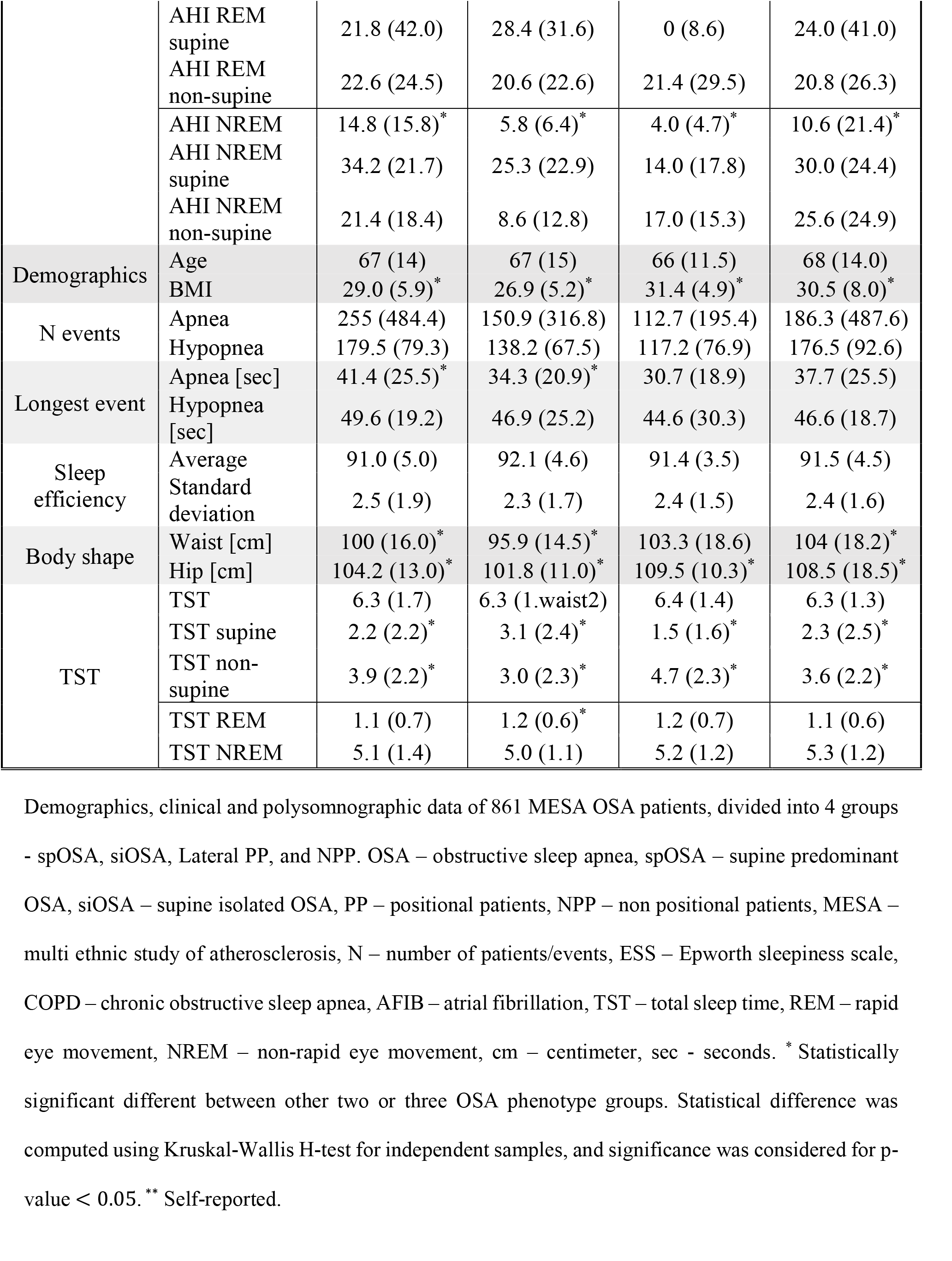
Demographics, clinical and polysomnographic data per Positional groups.

**Table 3:** Positional OSA data according to Ethnic group.

| Ethnicity | N | OSA<br>N (%) | Supine<br>PP<br>N (%) | spOSA<br>N (%) | siOSA<br>N (%) | Lateral<br>PP<br>N (%) | NPP<br>OSA<br>N (%) |
| --- | --- | --- | --- | --- | --- | --- | --- |
| White,<br>Caucasian | 493 | 319<br>(64.7) | 218<br>(68.3) | 96<br>(30.1) | 122<br>(38.2) | 15<br>(4.7) | 86<br>(27.0)* |
| Chinese<br>American | 167 | 104<br>(62.3) | 76<br>(73.1)* | 34<br>(32.7) | 42<br>(40.4) | 1<br>(1.0) | 27<br>(26.0) |
| Black,<br>African<br>American | 367 | 233<br>(63.5) | 131<br>(56.2)* | 64<br>(27.5) | 67<br>(28.8)* | 12<br>(5.2) | 90<br>(38.6)* |
| Hispanic | 296 | 205<br>(69.3) | 125<br>(61.0) | 63<br>(30.7) | 62<br>(30.2) | 7<br>(3.4) | 73<br>(35.6) |

spOSA, siOSA, Lateral PP, and NPP OSA prevalence of each MESA's ethnicity. The percentages of the first three are calculated by conditioned N (e.g., spOSA N) out of the total number of patients with OSA of the relevant ethnicity. \* Statistically significant different in OSA phenotype incidence between other two or three examined ethnic groups. Statistical difference was computed using Kruskal-Wallis H-test for independent samples, and significance was considered for p-value < 0.05. OSA – obstructive sleep apnea, spOSA – supine predominant OSA, siOSA – supine isolated OSA, PP – positional patients, NPP – non positional patients, MESA – multi ethnic study of atherosclerosis.

For Lateral PP, the trend of positional AHI in REM, and the trend of positional AHI in NREM, was opposite to all the other groups, and it reflected the same effect of position on positional AHI in general, i.e., supine AHI was greater than non-supine AHI for Supine PP (e.g., for spOSA 39.9 (32.5)>9.8 (9.0)), and for Lateral PP, non-supine AHI was higher (13.7 (13.3)>3.8 (4.6)). That is, for Lateral PP, AHI REM non-supine > AHI REM supine, and AHI NREM non-supine > AHI REM supine (21.4 (29.5)>0 (8.6), and 17.0 (15.3)>14.0 (17.8), respectively). Moreover, almost no apnea or hypopnea events occurred in the supine posture during REM sleep for most of the Lateral PP (0 (8.6)). We did not find a statistical difference in the prevalence of hypertension between Lateral PP compared to the other groups. However, hypertension prevalence among siOSA was significantly lower than of all other groups (49.1% versus 59.5%-65.7%, p-value<0.05).

### Ethnicity analysis

Table 3 shows that Black-African American OSA patients had a prevalence of 28.8% siOSA. This was significantly lower than Chinese Americans and White-Caucasians which have a prevalence of 40.4% and 38.2% (p-value ≤ 0.036) respectively. spOSA incidence among Black-African Americans was the least frequent (27.5%), but this, however, did not shown statistical significance. Overall, Chinese American tended to be more Supine PP (73.1%) and less Lateral PP (1.0%) compared to other ethnic groups. The Black-African American patients had a lower proportion of Supine PP (56.2%) and a higher proportion of Lateral PP (5.2%) compared to other ethnic groups. These observations where statistically significant (p-value = 0.003) for Supine PP but not for Lateral PP. We also found that Black-African Americans have significantly higher prevalence of NPP than Chinese Americans and White-Caucasians (38.6% versus 26.0%, and 27.0%, respectively, p-value ≤ 0.004) and that White-Caucasians are also statistically significant less NPP than Hispanics (27.0% vs. 35.6%, p-value = 0.036).

## Discussion

The results of this study highlighted two important novel findings: we present for the first time, in this relatively large group of OSA patients, the PP characteristics for multiple ethnic groups, and we report the existence of a new phenotype of OSA PP namely, the Lateral PP and provide demographic as well as polysomnographic characteristics for this group. The prevalence of Supine PP among OSA severity groups were: 69.1% for mild, 62.2% for moderate, and 52.4% for severe OSA. These results are in line with the results in the literature showing that Supine PP are mainly dominant in the less severe groups of patients with OSA^14^. Moreover, our findings on Supine PP and siOSA prevalence among OSA in MESA were also aligned with previous reports^6,14,^ Overall, up to 34.0% of all OSA individuals were siOSA, and by using positional therapy (PT) efficiently, they could resolve their OSA condition. In addition, another 29.8% of all the patients with OSA were spOSA, and therefore could have their OSA condition remarkably improved by using effectively this behavioral therapy.

### Characteristics of OSA PP according to ethnicity

We found that Chinese American had a higher prevalence of Supine PP, whereas Black-African American patients had significantly less Supine PP, as compared to other ethnic groups. In addition, we also found that Black-African Americans had significantly higher prevalence of NPP than Chinese Americans, and White-Caucasians. Also, compared to Hispanics, White-Caucasians were significantly less NPP. The relatively significantly low prevalence of Supine PP and high prevalence of NPP among Black-African Americans might be partially explained by some clinical, demographic, and polysomnographic differences with respect to the other ethnic groups (Table 4). Black-African Americans in MESA had significantly higher BMI, and waist circumference than White-Caucasian, and Chinese American. Black-African American suffer significantly more from hypertension and had a more severe daytime sleepiness level as assessed by the ESS score compared to all other ethnic groups. They had lower supine AHI, as compared to Chinese American and Hispanic. Contrary, AHI in REM sleep for Black-African American was significantly higher than White-Caucasians, and Chinese Americans.

**Table 4:** Demographics, clinical, and polysomnographic information of patients with OSA according to ethnic group.

| Category | Variable | White,<br>Caucasian | Chinese<br>American | Black,<br>African<br>American | Hispanic |
| --- | --- | --- | --- | --- | --- |
| N (%) |  |  |  |  |  |
|  | N | 319 | 104 | 233 | 205 |
| Gender | Male | 170 (53.3) | 65 (62.5) | 112 (48.1) | 101 (49.3) |
| Cigarette | Never | 133 (41.7) | 74 (71.1)* | 97 (41.6) | 96 (46.8) |
|  | Former | 170 (53.3) | 24 (23.1)* | 113 (48.5) | 94 (45.8) |
|  | Current | 15 (4.7) | 6 (5.8) | 22 (9.4) | 14 (6.8) |
|  | None | 0 (0.3) | 0 (0) | 1 (0.4) | 1 (0.3) |
| Hypertension | No | 152 (47.6) | 58 (55.8)* | 61 (26.2)* | 89 (43.4)* |
|  | Yes | 166 (52.0) | 46 (44.2)* | 172 (73.8)* | 116 (56.6)* |
|  | None | 1 (0.3) | 0 (0) | 0 (0) | 0 (0) |
| Asthma | No | 312 (97.8) | 101 (97.1) | 220 (94.4) | 280 (94.6) |
|  | Yes | 6 (1.9) | 3 (2.9) | 13 (5.6) | 15 (5.1) |
|  | Don't know | 0 (0) | 0 (0) | 0 (0) | 1 (0.3) |
|  | None | 1 (0.3) | 0 (0) | 0 (0) | 0 (0) |
| ESS | Normal (0-10) | 280 (87.8)* | 90 (86.5)* | 182 (78.1)* | 260 (87.8) |
|  | Mild (11-14) | 25 (7.8)* | 11 (10.6)* | 35 (15.0)* | 25 (8.5) |
|  | Moderate (15-17) | 10 (3.1)* | 0 (0)* | 10 (4.3)* | 6 (2) |
|  | Severe (18-24) | 2 (0.6)* | 1 (1.0)* | 4 (1.7)* | 4 (1.4) |
|  | None | 2 (0.6) | 2 (1.9) | 3 (1.3) | 1 (0.3) |
| Emphysema<br>or COPD** | No | 483 (98) | 103 (99.0) | 227 (97.4) | 202 (98.5) |
|  | Yes | 9 (1.8) | 1 (1.0) | 5 (2.1) | 2 (1.0) |
|  | Don't know | 0 (0) | 0 (0) | 1 (0.4) | 1 (0.5) |
|  | None | 1 (0.2) | 0 (0) | 0 (0) | 0 (0) |
| AFIB | No | 312 (97.8) | 166 (99.4) | 103 (99.0) | 203 (99.0) |
|  | Yes | 5 (1.6) | 1 (0.6) | 1 (0.4) | 1 (0.5) |
|  | None | 3 (0.6) | 0 (0) | 0 (0) | 1 (0.5) |
| MEDIAN (IQR) |  |  |  |  |  |
| AHI | AHI | 13.0 (14.6)* | 15.8 (15.9) | 14.5 (14.5) | 15.8 (18.7) |
|  | AHI supine | 23.1 (29.0) | 28.0 (31.5) | 20.0 (26.7)* | 25.0 (28.7) |
|  | AHI non-supine | 6.6 (12.1)* | 7.1 (12.3) | 8.9 (13.4) | 8.7 (13.1) |
|  | AHI REM | 3.9 (4.5)* | 3.8 (3.6)* | 5.1 (4.9)* | 5.7 (5.5)* |
|  | AHI REM supine | 18.7 (39.7) | 23.4 (42.4) | 28.6 (33.3) | 26.6 (33.3) |
|  | AHI REM non-supine | 21.4 (25.2) | 18.0 (19.4) | 22.2 (26.0) | 21.8 (24.2) |
|  | AHI NREM | 9.0 (12.0) | 12.0 (16.9) | 8.8 (13.4) | 9.3 (14.8) |
|  | AHI NREM supine | 29.1 (25.1) | 30.7 (23.1) | 27.1 (23.2) | 31.0 (24.3) |
|  | AHI NREM non-supine | 17.9 (22.1) | 16.6 (18.1) | 16.9 (18.9) | 17.8 (25.5) |
| Demographics | Age | 68 (15.0) | 64.5 (14.0) | 68.0 (14.0) | 67 (15.0) |
|  | BMI | 27.7 (6.3)* | 24.4 (3.8)* | 30.4 (7.1)* | 30.5 (6.7)* |
| N events | Apnea | 172.6 (379.0) | 215.0 (466.5) | 171.4 (419.2) | 214.0 (423.9) |
|  | Hypopnea | 160.6 (82.4) | 143.1 (80.4) | 154.9 (88.8) | 166.4 (86.2) |
| Longest event | Apnea [sec] | 35.2 (23.9)* | 37.7 (29.2)* | 34.4 (20.0)* | 41.8 (28.4)* |
|  | Hypopnea [sec] | 47.3 (24.0) | 52.3 (25.7)* | 44.8 (18.4)* | 47.5 (18.8)* |
| Sleep efficiency | Average | 92.4 (4.3)* | 91.5 (4.4) | 90.6 (4.8) | 91.4 (4.8) |
|  | Standard deviation | 2.3 (1.5)* | 2.6 (1.6) | 2.8 (1.9) | 2.4 (1.7)* |
| Body shape | Waist [cm] | 99.5 (17.4)* | 89.2 (9.3)* | 102.8 (16.7)* | 103.0 (14.0)* |
|  | Hip [cm] | 104.6 (12.8)* | 95.7 (7.2)* | 109.0 (16.8)* | 105.6 (13.8)* |
| TST | TST | 6.5 (1.5)* | 6.2 (1.4) | 6.1 (1.5) | 6.3 (1.3) |
|  | TST supine | 2.4 (2.4) | 2.7 (2.0) | 2.7 (2.4) | 2.7 (2.4) |
|  | TST non-supine | 3.8 (2.4)* | 3.3 (1.8) | 3.2 (2.4) | 3.6 (2.4) |
|  | TST REM | 1.2 (0.6) | 1.0 (0.6)* | 1.1 (0.7) | 1.2 (0.6) |
|  | TST NREM | 5.2 (1.3) | 5.0 (1.2) | 5.3 (1.2) | 5.1 (1.2) |

Demographics, clinical, and polysomnographic information of 861 MESA OSA patients, divided into 4 ethnic groups – White-Caucasian, Chinese American, Black-African American, and Hispanic. Interesting findings are marked in yellow. N – number of patients/events, IQR - interquartile range, ESS – Epworth sleepiness scale, COPD – chronic obstructive sleep apnea, AFIB – atrial fibrillation, TST – total sleep time, REM – rapid eye movement, NREM – Non-rapid eye movement, cm – centimeter, sec - seconds. \* Statistically significant different between other two or three examined ethnic groups. Statistical difference was computed using Kruskal-Wallis H-test for independent samples, and significance was considered for p-value < 0.05. \*\* Self-reported.

It has already been reported many times that Supine PP are OSA patients with a less severe disease compared to NPP. Therefore, the NPP condition is, overall, more severe, with a worst sleep quality and with more daytime sleepiness compared to PP. Accordingly, the data obtained in the present study suggest that since Chinese American have a higher prevalence of PP, it is the group of OSA patients that is supposed to have an overall less severe disease burden. Similarly, since Black-African Americans have a high prevalence of NPP, the overall severity of OSA in this group is expected to be higher. However, these positional prevalence differences were not reflected by the AHI across ethnic groups (Table 4). These results suggest that the differences in positional prevalence found between ethnic groups in this study, although statistically significant, are not sufficient enough to translate into establishing AHI severity differences.

Black-African Americans in the MESA population also had features that are consistent with the notion of a group of OSA patients with a severe disease. For example; they have a significantly higher BMI compared to White-Caucasian, and Chinese American and it is well known that in general, increased BMI is correlated with a higher OSA severity^8^. The body characteristics of these patients also support the higher BMI, with waist and hip circumference significantly higher than all the other ethnic groups. In addition, from the clinical perspective, Black-African American had a high prevalence of hypertension, and also were subjectively sleepier during daytime, as compared to all three other ethnic groups. Hypertension and daytime sleepiness are also two clinical conditions that are more commonly seen in OSA patients with a severe disease. Therefore, all these previous data support the notion that Black-African American would be an OSA group with a supposedly more severe form of the disease. Already in 1995, S. Ancoli-Israel et al.^15^ found that more African-Americans had severe OSA with a relative risk=2.13 compared to Caucasians, and this result was confirmed further after a logistic regression analysis where race was associated with the presence of severe OSA (RDI (old AHI) ≥ 30/h) independently of age, sex, and BMI. The mean RDI for those African-Americans with severe OSA was significantly higher than that for Caucasians. One reason for the high severity of the disease among these OSA patients may be the late diagnosis and underrepresented in sleep disorders units of this group compared to the other ethnic groups^16^. Nevertheless, in our dataset Black African American patients did not have a more severe disease (expressed by the AHI) than the other ethnic groups. However, it was noticed that they did have the highest REM AHI in the supine posture.

Asians OSA patients had a high Supine PP prevalence (around 75%). The high prevalence in this group was previously reported by Mo et al^10^ A possible explanation is the short cranial base and retrognathia that are more common among Asians, and may lead to an airway structure that tends to collapse in supine position^17^. Caucasians’ Supine PP prevalence in the literature has been reported to be in the range 26.7-55.9%^17^. There is a lack of research regarding other ethnic groups in the context of Supine PP. According to our findings, the Chinese American ethnic group is most affected by the sleeping position with 74.1% of all patients having either Supine or Lateral PP, as compared to 61.4-73% for the other ethnic groups.

### Demographic and polysomnographic characteristics of Lateral PP

Our analysis highlighted a novel phenotype of PP we coined “Lateral PP”, that included patients with OSA having a non-supine AHI more than double the supine AHI. This was a small group of patients (4.1%) among the 861 OSA patients. Lateral PP were mainly obese, female, and with mild-moderate OSA. Chinese American was the group of OSA patients with the lowest prevalence of Lateral PP compared to the other ethnicity groups. About a third of Lateral PP slept less than 1-hour in the supine position and therefore, it is possible that the reason they did not show apneas/hypopneas in the supine posture is the limited time slept in this posture. This is critical issue that could be solved by having these patients sleep several nights in the lab in order to investigate whether the results stay consistent. One possibility is that those are supine PP OSA patients that spontaneously avoid the supine posture since they know, based on their experience, that this sleeping position worsen their OSA condition. Nevertheless, another possibility is that this could also be a specific group of OSA patients for whom the condition is truly worst in the non-supine postures compared to the supine position. Yet, it is important to notice that the majority of the Lateral PP (71.4%) slept more than 1 hour in supine, which for them, the reason of limited sleep time in supine is less likely. We do not know if these patients had some unique cephalometric characteristics, and this is certainly also a topic for future research. It is important to note that the non-supine AHI of Lateral PP was significantly higher than the non-supine AHI of Supine PP, and most of the breathing abnormalities occurred during REM sleep, a known characteristic of many women with OSA^18^. All these observations require confirmation from the study of other datasets of patients with OSA and ideally the study of those Lateral PP over several nights to investigate the night to night prevalence variability of this novel positional phenotype.

If this new PP phenotype, named “Lateral PP” is confirmed, it may have clinical consequences on the treatment recommendations for this group of patients. Lateral PP would be advised to adopt the supine posture and avoid the lateral position i.e. the opposite recommendation given to Supine PP. Indeed, PT could be an appropriate therapy for Lateral PP. Nevertheless, how effective PT would be in Lateral PP stayed to be researched. We know that compliance with PT is a challenge. During many years the Tennis Ball Technique (TBT) was the popular PT for avoiding the supine posture during sleep but this therapy was uncomfortable and the compliance was low^19^. Nevertheless, during the last years a new generation of devices for PT which provide a subtle vibrating stimulus that prevents patients adopting the supine position has been used successfully by many OSA Supine PP and the results are good for short-term use but still limited for long term use^19,20^.

### Limitations

The study considered only one-night PSG examination, and it is known that consecutive night PSG study can change the prevalence of PP and NPP^21^. Moreover, another fact that might influence the results and the prevalence of OSA and its phenotypes (e.g., Supine PP, Lateral PP) is that we used apnea and hypopneas’ definitions that are not align with the latest AASM 2012 definitions^22^. We chose to use the most common definition for PP (Cartwright’s definition), but using other definition for OSA PP could provide different results. It should be mentioned that we cannot surely refute that Lateral PP phenotype is not a consequence of different positional sleep time, as we rejected KS’s null hypothesis for supine sleep time, for lateral sleep time, and for TST_supine_/TST, meaning that the distribution of those statistics for Lateral PP and supine PP were different. This requires further investigation.

## Conclusion

In this study we describe for the first time, demographic and polysomnographic characteristics of positional patients (PP), over different ethnic groups in a relatively large population of OSA patients. In addition, we report a novel OSA PP phenotype, namely Lateral PP, and characterize those patients demographically and polysomnographic-wise. Future research will need to confirm the results of this preliminary report.

## Data Availability

All data produced are available online at National Sleep Research Resource (NSRR)

https://sleepdata.org/

## Abbreviations

AHI: Apnea-hypopnea index
IQR: Interquartile range
IRB: Institutional review board
KS: Kolmogorov-Smirnov
KW: Kruskal-Wallis
MESA: Multi-ethnic study of Atherosclerosis
NHLBI: National Heart, Lung and Blood Institute
NSRR: National Sleep Research Resources
ODI: Oxygen desaturation index
OSA: Obstructive sleep apnea
POSA: Positional obstructive sleep apnea
PSG: Polysomnography
PT: Positional Therapy
RDI: Respiratory distribution index
TBT: Tennis ball technique
TST: Total sleep time
liOSA: Lateral-isolated obstructive sleep apnea
lpOSA: Lateral-predominant obstructive sleep apnea
siOSA: Supine-isolated obstructive sleep apnea
spOSA: Supine-predominant obstructive sleep apnea

## Acknowledgement

The Multi-Ethnic Study of Atherosclerosis (MESA) Sleep Ancillary study was funded by NIH-NHLBI Association of Sleep Disorders with Cardiovascular Health Across Ethnic Groups (RO1 HL098433). MESA is supported by NHLBI funded contracts HHSN268201500003I, N01-HC-95159, N01-HC-95160, N01-HC-95161, N01-HC-95162, N01-HC-95163, N01-HC-95164, N01-HC-95165, N01-HC-95166, N01-HC-95167, N01-HC-95168 and N01-HC-95169 from the National Heart, Lung, and Blood Institute, and by cooperative agreements UL1-TR-000040, UL1-TR-001079, and UL1-TR-001420 funded by NCATS. The National Sleep Research Resource was supported by the National Heart, Lung, and Blood Institute (R24 HL114473, 75N92019R002).

## Supplements

### Supplement 1 – MESA events’ definitions

The MESA group used the following cited definitions according to the PSG Scoring Manual they have published under https://sleepdata.org/datasets/mesa/pages/polysomnography-introduction.md in NSRR website –

#### “Obstructive Apneas

will be identified if the amplitude (peak to trough) of the airflow signal is flat or nearly flat. This is noted when amplitude of airflow decreases below at least 90% of the amplitude of “baseline” breathing, i.e. to < 10% of the baseline for 90% of the event duration, (identified during a period of regular breathing with stable oxygen levels), and if this change lasts for > 10 s. When thermistor is missing or uninterpretable, the nasal pressure signal (cannula flow) can be used as an alternative signal for scoring Obstructive Apnea.”

#### “Central Apneas

will be noted if no displacement is noted on both chest and the abdominal inductance channels. Otherwise, events will be noted as “obstructive”. Central events cannot be designated if either or both band data are missing or uninterpretable.”

#### “<u>AASM Alternative Hypopneas</u> (50% reduction of amplitude)

Will be identified if > 50% reduction of amplitude is visualized on both belts (thoracic and abdominal). Alternatively, an AASM Alternative hypopnea may be marked if a clear > 50% reduction is seen on a good nasal pressure signal. Specifically, when amplitude changes are seen only in the nasal pressure with little or no changes in the belt channels, the event will not be marked. These events are marked independent of whether any associated desaturation is linked with the event.”

#### “<u>AASM Recommended Hypopneas</u> (30% reduction of amplitude)

will be identified if the amplitude of the thorax and abdomen signals decrease by at least (approximately) 30% of the amplitude of “baseline”, i.e. to < 70% of baseline, (identified during a period of regular breathing with stable oxygen levels), if this change lasts for > 10 s, and if the event does not meet the criteria for the AASM Alternative hypopnea. These events are marked independent of whether any associated desaturation is linked with the event.”

Where AASM refers to AASM 2007.

Based on these definitions, a pre-calculated table, supplied by MESA group, has been generated, with various definitions for each event/parameter. Finally, based on previous MESA studies^S1,S2,S3,^ we used the following parameters and definitions –

**AHI** = (oahi4_rem5 · time in REM + oahi4_nrem5 · time in NREM) / TST Where

- oahi4_rem5 = Obstructive Apnea (all desaturations); 30% and 50% hypopnea (4% desaturations) index in REM.
- oahi4_nrem5 = Obstructive Apnea (all desaturations); 30% and 50% hypopnea (4% desaturation) index in NREM.

**AHI**_**supine**_ = oahi4_sup5 = Obstructive Apnea (all desaturations); 30% and 50% hypopnea (4% desaturation) index supine.

**AHI**_**non-supine**_ = oahi4_nsup5 = Obstructive Apnea (all desaturations); 30% and 50% hypopnea (4% desaturation) index non-supine.

**N Apneas** = oai0p5 * TST + cai0p5 * TST Where:

- oai0p5 = Calculated – Obstructive apnea index all desats
- cai0p5 = Calculated – Central apnea index all desats

**N Hypopneas** = hunrop5 + hurop5 + hurbp5 + hurnbp5 + rdirbp5 + rdirop5 + rdinbp5 + rdinop5 Where:

- hunrop5 = # AASM alternative hypopnea per hour (NREM, OTHER, all desaturations)
- hurop5 = # AASM alternative hypopnea per hour (REM, OTHER, all desaturations)
- hurbp5 = # AASM alternative hypopnea per hour (REM, BACK, all desaturations)
- hurnbp5 = # AASM alternative hypopnea per hour (NREM, BACK, all desaturations)
- rdinop5= # AASM recommended hypopnea per hour (NREM, OTHER, all desaturations)
- rdirop5 = # AASM recommended hypopnea per hour (REM, OTHER, all desaturations)
- rdirbp5 = # AASM recommended hypopnea per hour (REM, BACK, all desaturations)
- rdinbp5 = # AASM recommended hypopnea per hour (NREM, BACK, all desaturations)

### Supplement 2 – Additional figures

**Figure S1:**
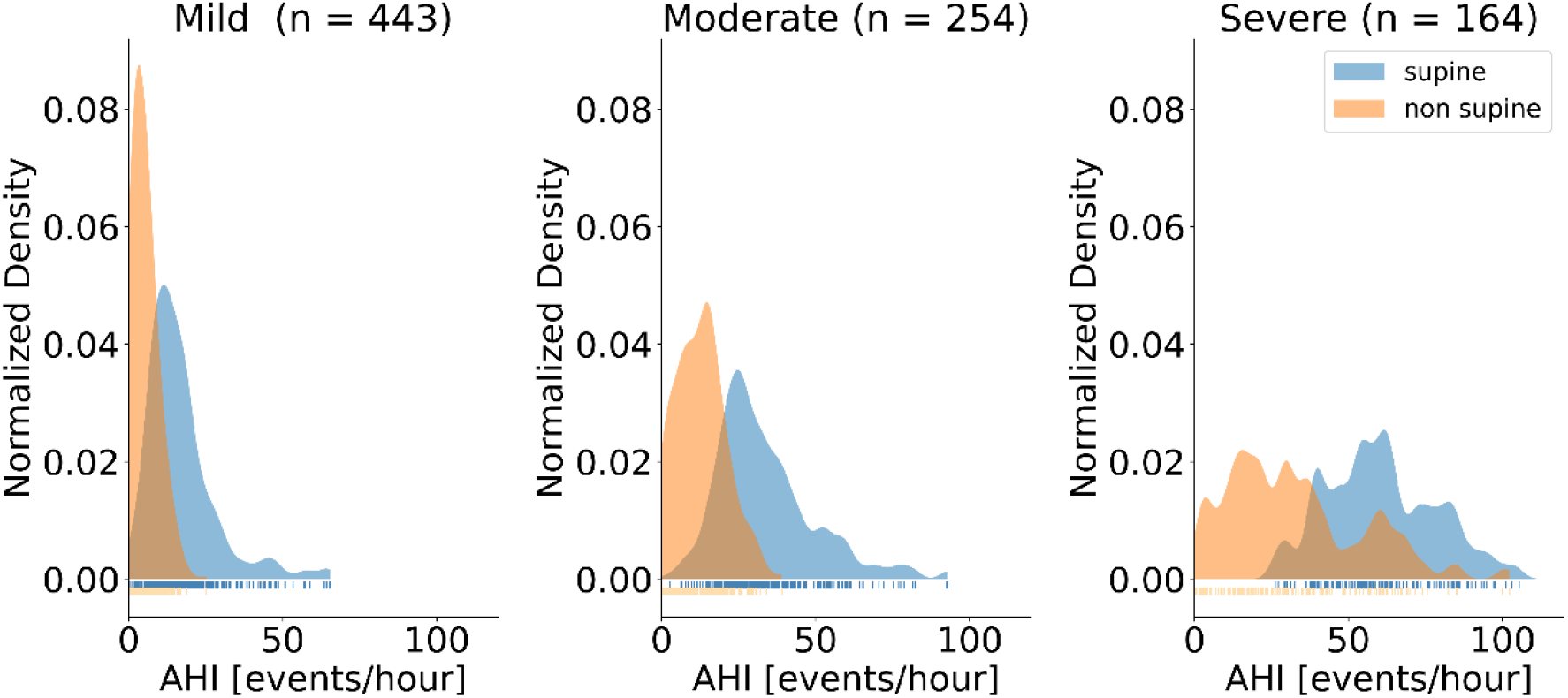
Apnea-hypopnea index distribution. Kernel density estimation (KDE) of MESA AHI divided by OSA severity (mild, moderate, and severe), and position (supine, and non-supine). AHI – apnea-hypopnea index, OSA – obstructive sleep apnea.

**Figure S2:**
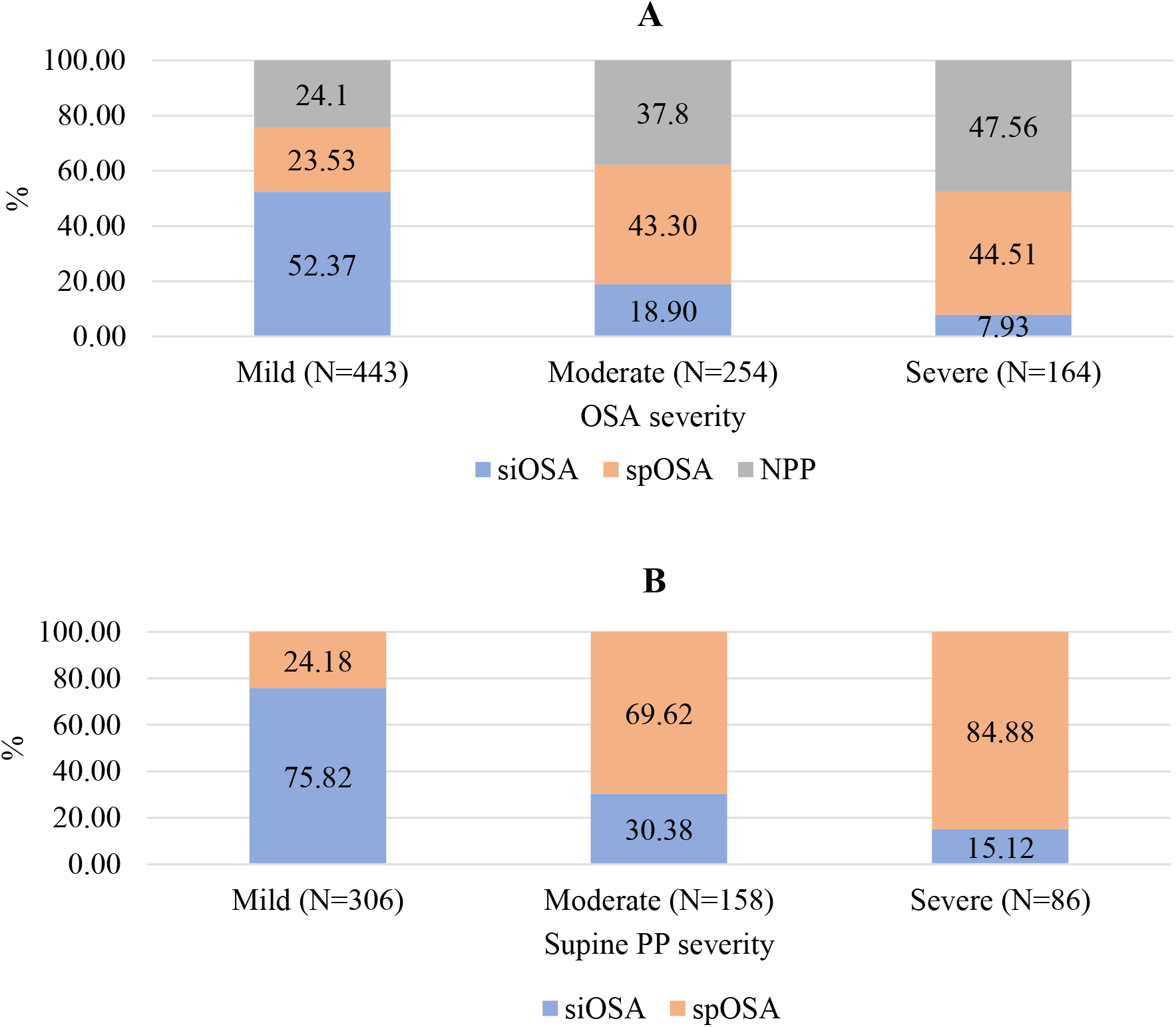
OSA phenotypes and Supine PP phenotypes prevalence among different severities. Stacked bar-plot of (A) Supine Predominant OSA (spOSA), Supine Isolated OSA (siOSA), and Non Positional Patients (NPP) prevalence percentages among different OSA severity categories, and (B) spOSA, and siOSA incidence out of Supine PP (Cartwright definition^9^). To calculate Supine PP percentages according Cartwright definition, sum siOSA and spOSA incidences. OSA – obstructive sleep apnea, spOSA – supine predominant OSA, siOSA – supine isolated OSA, PP – positional patients, NPP – non positional patients, MESA – multi ethnic study of atherosclerosis N – number of patients in the specific severity.

**Figure S3:**
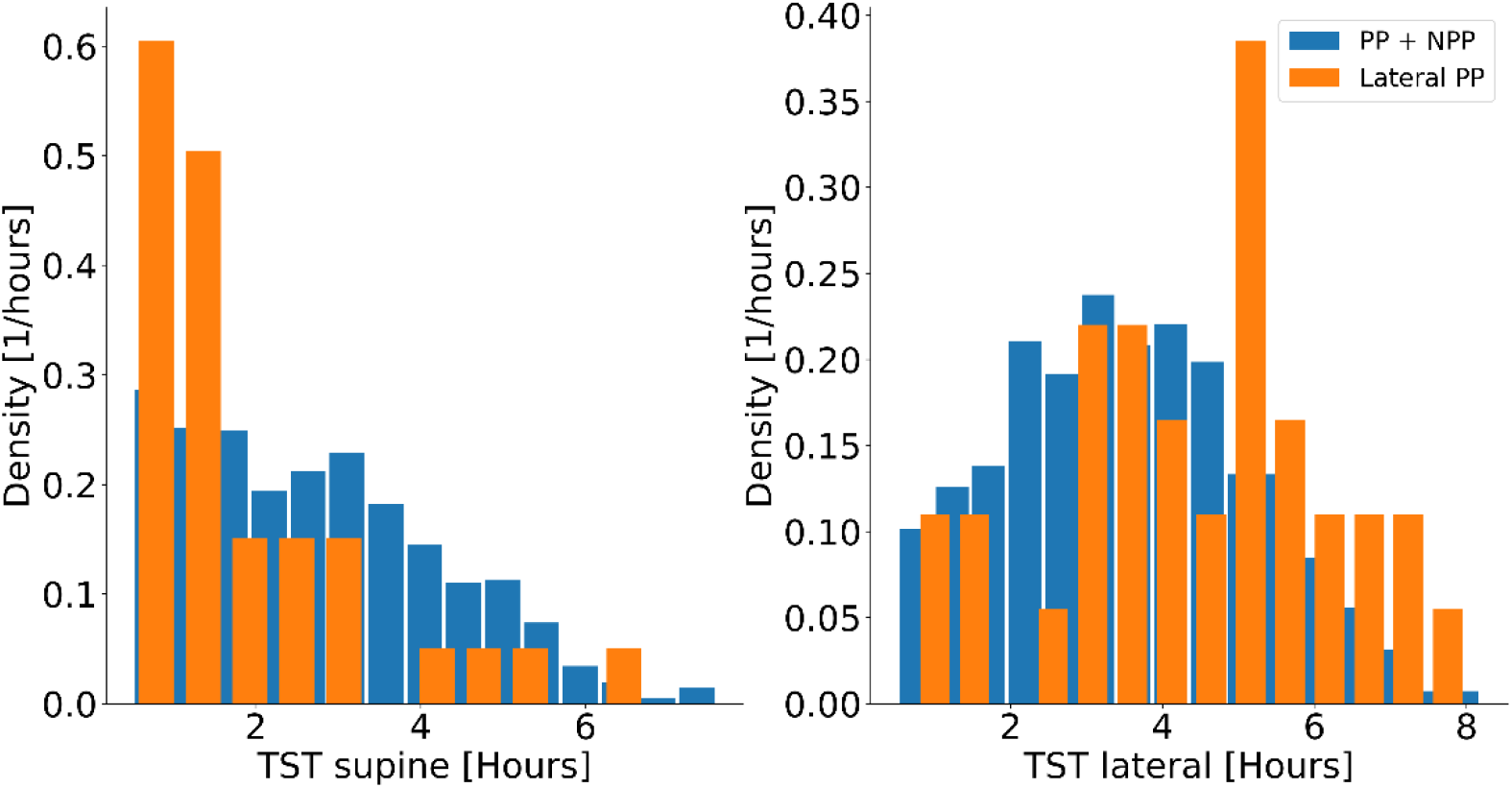
Positional total sleep time distributions of Lateral PP versus rest patients with OSA. Histogram of TST_supine_, and TST_non-supine_ (hour by hour) for Lateral PP versus all the rest MESA relevant patients (i.e., supine PP, NPP). PP – positional patients, NPP – non positional patients, TST – total sleep time.

**Figure S4:**
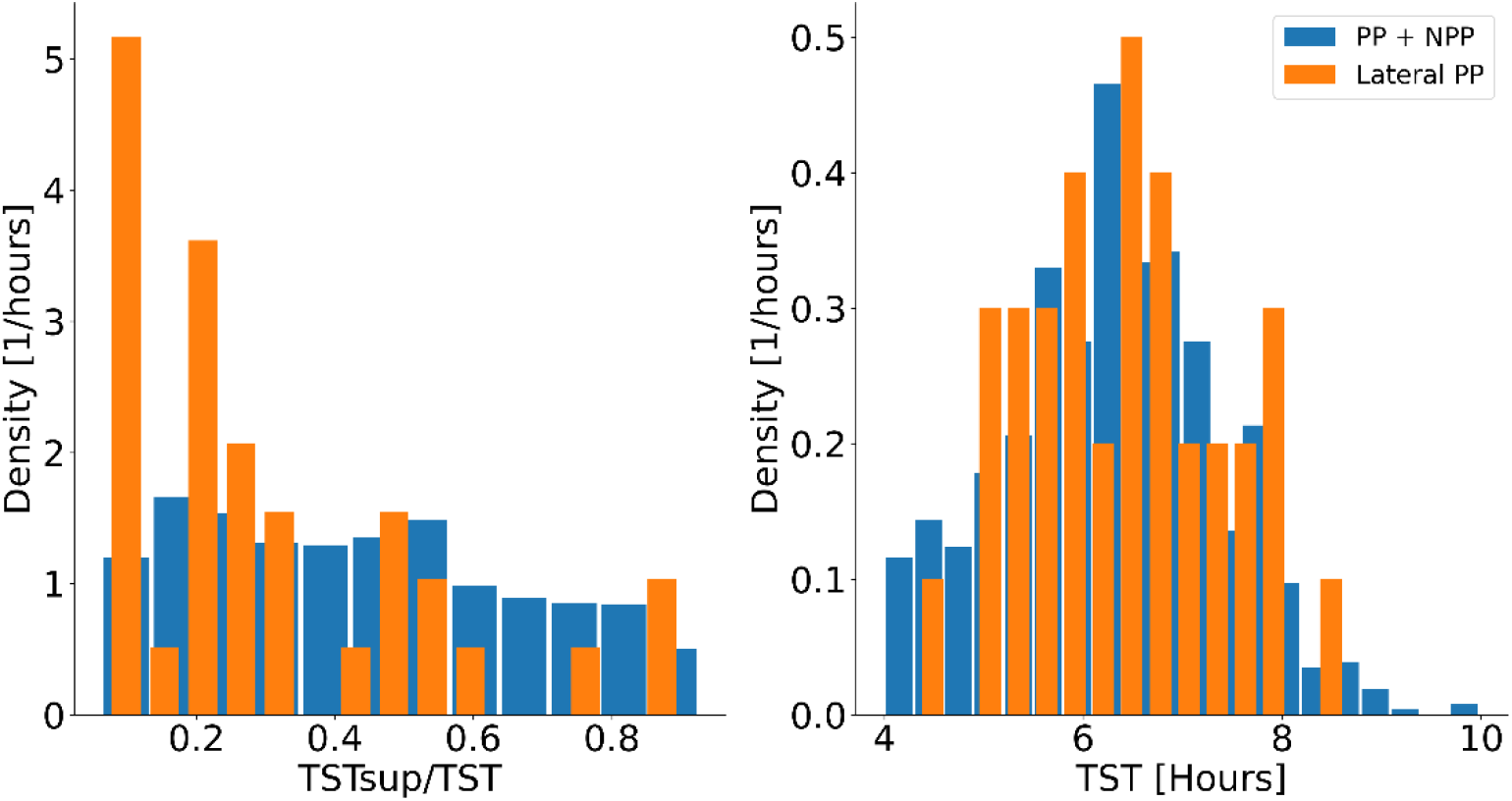
Total sleep time (TST) and proportional supine TST distributions of Lateral PP versus rest patients with OSA. Histograms of *TST*_*supine*_/*TST*, and TST (hour by hour) for Lateral PP versus all the rest MESA relevant patients (i.e., supine PP, NPP). PP – positional patients, NPP – non positional patients, TST – total sleep time.

### Supplement 3 – Additional tables

**Table S1:** TST and positional TST of each patient of the group of patients with OSA with Lateral PP.

| Patient # | OSA Group | TST supine | TST lateral | TST prone | TST | AHI supine | AHI non-supine | AHI |
| --- | --- | --- | --- | --- | --- | --- | --- | --- |
| 74 | lpOSA | 3.16 | 3.61 | 0 | 6.77 | 6.63 | 14.93 | 11.1 |
| 301 | liOSA | 1.22 | 5.22 | 0 | 6.43 | 2.47 | 13.99 | 11.8 |
| 465 | lpOSA | 1.05 | 4.55 | 0 | 5.60 | 6.67 | 21.32 | 18.6 |
| 920 | liOSA | 1.60 | 3.68 | 0 | 5.28 | 3.75 | 12.22 | 9.7 |
| 1090 | lpOSA | 0.93 | 6.90 | 0 | 7.83 | 47.14 | 99.86 | 93.6 |
| 1284 | lpOSA | 5.09 | 0.76 | 0 | 5.85 | 27.25 | 61.30 | 31.8 |
| 1476 | liOSA | 0.52 | 5.57 | 0 | 6.08 | 1.94 | 6.65 | 6.2 |
| 1698 | liOSA | 1.30 | 3.92 | 0 | 5.22 | 2.31 | 10.42 | 8.4 |
| 1907 | liOSA | 0.91 | 6.99 | 0.15 | 7.90 | 2.18 | 13.71 | 12.4 |
| 1973 | liOSA | 0.68 | 5.32 | 0 | 6.00 | 0.00 | 9.59 | 8.5 |
| 1995 | liOSA | 2.42 | 3.13 | 0 | 5.55 | 2.07 | 7.98 | 5.4 |
| 2339 | liOSA | 0.75 | 6.14 | 0 | 6.88 | 2.67 | 21.46 | 19.5 |
| 2349 | lpOSA | 2.68 | 3.01 | 1.23 | 5.68 | 7.83 | 39.12 | 24.5 |
| 2523 | lpOSA | 2.67 | 1.68 | 0 | 4.35 | 6.75 | 19.01 | 11.5 |
| 2539 | lpOSA | 4.12 | 3.34 | 0 | 7.47 | 10.65 | 29.85 | 19.3 |
| 2548 | lpOSA | 1.48 | 5.32 | 0 | 6.80 | 6.07 | 12.56 | 11.2 |
| 2739 | liOSA | 0.53 | 4.67 | 0 | 5.20 | 3.75 | 8.14 | 7.7 |
| 2802 | liOSA | 0.70 | 6.43 | 0.47 | 7.13 | 1.43 | 6.20 | 5.7 |
| 2892 | lpOSA | 4.97 | 1.41 | 0.01 | 6.38 | 11.44 | 31.06 | 15.8 |
| 3836 | liOSA | 1.50 | 5.77 | 0 | 7.27 | 4.00 | 15.95 | 13.5 |
| 4053 | lpOSA | 1.23 | 5.15 | 1.3 | 6.38 | 6.49 | 17.42 | 15.4 |
| 4334 | lpOSA | 6.75 | 0.75 | 0 | 7.50 | 11.67 | 25.33 | 13.1 |
| 4396 | liOSA | 1.55 | 3.94 | 2.46 | 5.48 | 0.00 | 12.66 | 9.1 |
| 4512 | liOSA | 2.08 | 4.27 | 0 | 6.35 | 2.40 | 6.80 | 5.4 |
| 4849 | lpOSA | 2.97 | 3.32 | 0.22 | 6.28 | 14.49 | 30.75 | 23.1 |
| 5393 | liOSA | 1.58 | 6.47 | 0 | 8.05 | 1.89 | 6.96 | 6.0 |
| 5832 | liOSA | 0.58 | 5.22 | 0 | 5.80 | 0.00 | 16.05 | 14.5 |
| 5882 | liOSA | 1.05 | 5.85 | 0 | 6.90 | 2.86 | 6.14 | 5.7 |
| 5887 | liOSA | 1.90 | 5.07 | 0.41 | 6.97 | 2.11 | 6.91 | 5.6 |
| 6002 | liOSA | 1.68 | 3.47 | 1.78 | 5.15 | 4.16 | 15.00 | 11.5 |
| 6003 | lpOSA | 0.58 | 7.05 | 0 | 7.63 | 8.57 | 22.98 | 21.9 |
| 6238 | liOSA | 1.48 | 3.62 | 0.77 | 5.10 | 0.67 | 8.02 | 5.9 |
| 6291 | liOSA | 0.62 | 8.02 | 0.11 | 8.63 | 3.24 | 8.34 | 8.0 |
| 6471 | liOSA | 1.34 | 5.12 | 0 | 6.47 | 1.48 | 6.99 | 5.9 |
| 6619 | lpOSA | 3.32 | 2.70 | 0 | 6.02 | 5.13 | 12.96 | 8.6 |

In green are liOSA group, in bright yellow are Lateral PP with $TST_{supine}$ or $TST_{lateral} \leq 1$ hour, and in pink are patients with $TST_{prone} > 0.5$ hour. OSA – obstructive sleep apnea, lpOSA
– lateral predominant OSA, liOSA –lateral isolated OSA, PP – positional patients, NPP – non positional patients.

